# Maternal Willingness to Participate in Research Involving Neuroimaging, Biological Sample Collection, and Data Storage: Towards a Multicentre Neurodevelopmental Research in a low-resource setting

**DOI:** 10.64898/2026.02.22.26346849

**Authors:** Albert Dayor Piersson, Christiana Amartey, Sarah Teiko Quartei, Klenam Dzefi-Tettey, Promise Emmanuel Sefogah, Aquel Rene Lopez

## Abstract

**Background:** Maternal participation in neurodevelopmental research involving neuroimaging and diverse biological samples is essential for understanding prenatal influences on early brain development, yet willingness in low-resource settings remains underexplored.

**Method:** We surveyed 300 mothers using a structured questionnaire to assess willingness to undergo brain health testing (with a focus on electroencephalography [EEG] and brain magnetic resonance imaging [MRI]), provide biological samples (blood, stool, urine, breast milk, placenta, amniotic fluid, vaginal/nasal fluid, saliva, tears), and consent to 10-year storage. Responses were analysed to examine associations between maternal sociodemographic factors and willingness to consent for each research component.

**Results:** Ninety-two percent of participants expressed willingness for brain health testing, including ∼82% and ∼88% interest in EEG and MRI, respectively, even for untreatable conditions. Self-reported histories of foetal defects (5.3%) and birth defects (7.3%) were notably low. Biospecimen acceptance was highest (>95%) for routine samples (blood, stool, urine) but significantly low for sensitive specimens (breast milk, placenta, amniotic fluid: 51–55%) including (vaginal fluid, saliva, tears: 16–47%). Higher levels of maternal education consistently predicted consent across modalities, while being in a relationship increased willingness for stool, urine, placenta, amniotic fluid, MRI, and EEG. Low income reduced uptake for placenta, amniotic fluid, MRI, and EEG. Only 48% consented to 10-year storage of images and samples for future research.

**Conclusion:** This study demonstrates high maternal willingness for neurodevelopmental research involving brain health testing and routine biospecimens in a low-resource setting. The findings highlight the feasibility of such protocols in a low-resource setting while exposing persistent inequities that risk underrepresenting disadvantaged mothers in maternal-child brain research. Contextually tailored consent models and capacity-building initiatives will be essential to ensure equitable, sustainable engagement across diverse LMIC populations.

## 1. Introduction

During gestation, factors such as maternal nutrition, stress hormones, immune status, and exposure to environmental toxins can epigenetically and directly influence foetal neurogenesis, neuronal migration, synaptogenesis, and myelination [1–5]. Postnatally, maternal caregiving behaviours, breastfeeding, social support, and the provision of a stimulating environment continue to exert significant influence on synaptic plasticity of the child’s brain, the maturation of neural circuits [4] and long-term cognitive and behavioural outcomes. Understanding these prenatal and postnatal influences often requires advanced methodologies such as neuroimaging, cognitive testing, and testing of biological samples to elucidate how the biological, environmental, and behavioural influences shape the developing brain. Techniques such as magnetic resonance imaging (MRI) [6], electroencephalography (EEG) [7, 8], and eye tracking [9] may offer unprecedented insight into the structural, functional, and molecular processes shaping the developing brain.

Maternal biological samples provide complementary windows on prenatal exposures, molecular pathways, and maternal–foetal interactions shaping neurodevelopment from gestation into early life. Maternal blood captures micronutrient and toxicant profiles, with elevated manganese reportedly associated with poorer early childhood outcomes and higher selenium linked to increased autism spectrum disorder (ASD) or attention-deficit/hyperactivity disorder risk [10, 11]. The maternal gut microbiome, profiled via stool, appears to have a stronger influence on infant neurodevelopment than the infant’s own early microbiome, and maternal dysbiosis has been associated with increased risks of neurodevelopmental disorders [12, 13]. Urinary biomarkers of environmental toxicants (e.g., bisphenols, metals) have been linked to deficits in gross motor and problem-solving skills [14, 15]. Breast milk integrates maternal health, environmental exposures, and nutritional signalling, simultaneously supporting neurodevelopment and carrying potential neurotoxicants such as heavy metals [16, 17]. Placental tissue, central to foeto-maternal nutrient and gas exchange, endocrine function, and immune modulation, offers critical insight into in utero programming of brain development [18, 19]. Amniotic fluid, enriched in metabolites such as 3-hydroxybutyrate relative to maternal blood, provides a direct readout of foetal brain energy metabolism [20, 21]. The vaginal microbiome constitutes another potential causal pathway, with early evidence that vaginal microbiota transfer may safely modulate neurodevelopment [22, 23].

Additional non-invasive matrices extend this neurodevelopmental “biospecimen toolkit.” The nose–brain interface contributes to immune regulation and cerebrospinal fluid clearance, and is being targeted for intranasal neuropeptide delivery, including arginine vasopressin, in autism trials [24, 25]. Saliva, which broadly mirrors plasma composition, is emerging as a feasible biomarker source for ASD screening and etiological studies, facilitating recruitment of neurotypical controls [26, 27]. Maternal tear fluid, rich in proteins, lipids, metabolites, nucleic acids, and electrolytes, reflects systemic and ocular inflammation, and infant tears have been shown to activate cortical regions, including the lateral occipital and pre/postcentral gyri on functional MRI [28–31]. Prior work has mainly examined parental attitudes toward single procedures such as imaging or isolated biospecimen collection [32–36]. However, few studies have systematically explored factors shaping requires integrated neuroimaging and biological sampling to elucidate how maternal factors influence offspring brain development. This study addresses that gap by assessing maternal willingness to participate in such protocols, aiming to inform inclusive, culturally sensitive, and ethically robust strategies for sustained engagement from pregnancy through early childhood.

## Methods

### 2.1 Study Design

This study employed a cross-sectional design to assess maternal interest in and willingness to consent to participate in research that involves neuroimaging and the provision of biological samples for testing, geared towards multicentre neurodevelopmental research, with a focus on identifying the demographic and socioeconomic factors that influence consent.

### 2.2 Participants, Recruitment, and Sampling

We targeted 385 pregnant women and mothers of infants/young children using convenience sampling from antenatal clinics and postpartum units at a public tertiary facility in Ghana. Eligible participants were mothers aged ≥18 years, fluent in English or a local language with validated translations, capable of providing informed consent, and (for prenatal recruitment) carrying singleton pregnancies without known major foetal anomalies. We excluded those with severe cognitive impairment precluding consent or a history of substance abuse that could compromise data reliability.

The target sample size of 385 was determined using Cochran’s formula for survey research:

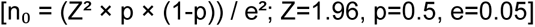

We received responses from 300 women.

### 2.3 Data Collection

Data was collected using a structured pretested questionnaire which included sociodemographic and (age, relationship status, educational attainment, monthly income [GH₵], health insurance subscription), and history of foetal or birth defect (Yes/No).

Maternal willingness was assessed on the questionnaire by presenting hypothetical scenarios where participants were to indicate their willingness. We adopted questions 1 and 2 from a previous study [37]:

1. “Would you take a test even if it provides information about a disease that cannot be prevented or treated?”
2. “Imagine a simple brain health test to learn about risk of developing a brain disease. Would you wish to take such a test?”. We complemented this question with the options of two breastmilk, nasal fluid, amniotic fluid, tears, saliva, vaginal fluid, stool, and placenta. Respondents were provided the options to select Yes, No, or Not sure.
3. We developed a question to assess willingness to allow the storage of your images/biological fluids to be stored for at least 10 years for future research. Respondents were provided the options to select Yes, No, or Not sure.

### 2.4 Data Analysis

All statistical analyses were performed using Real statistics. Age was reduced to young adults (18 – 30 years), middle-aged adults (31 – 40 years), and older adults (≥ 41 years). Relationship status was reduced to two variables as either “married” or “single” (widowed, divorced and single). We reduced highest education level to either “higher education” (professional training, higher national diploma, university) or “lower education” (none, primary school, junior high school, senior high school, vocational training). Then we stratified the income status as follows: **“**low Income**”** (GHc0, 0 – 999), “middle Income” (1,000 – 1,999 and 2,000 – 2,999), or **“**high Income**”** (3,000 – 3,999 and ≥4,000). Frequencies and percentages were used to describe categorical variables, while means and standard deviations (or medians and interquartile ranges) were used to describe continuous variables. Responses to the questions on willingness were dichotomised. Chi-square tests or independent t-tests were performed to examine associations between maternal sociodemographic factors and willingness to consent for each research component. Multivariable Logistic Regression models were constructed for each willingness outcome to identify independent predictors. Maternal sociodemographic and health factors were entered as independent variables. Odds Ratios (OR) and 95% Confidence Intervals (CI) were reported. Statistical significance was set at *p* <0.05.

### 2.5 Ethical Consideration

The study protocol was approved by the Institutional Review Board of the University of Cape Coast (UCCIRB/EXT/2022/30). All participants were provided with detailed information about the study objectives, procedures, potential risks, and benefits, and were given the opportunity to ask questions before participation. Informed written consent was obtained from all participants before enrolment, with consent documented through signed consent forms in accordance with the approved ethical protocol and the principles of the Declaration of Helsinki. To maintain confidentiality, data were anonymised and used exclusively for research purposes. Emphasis was placed on the voluntary nature of participation and the right to withdraw at any time without penalty.

## Results

Table 1 shows the descriptive statistics of participants’ sociodemographic characteristics.

**Table 1.** Sociodemographic characteristics of participants.

| Respondents | n | % of total |
| --- | --- | --- |
| Age range (years) |  |  |
| 18 – 30 | 205 | 68.3 |
| 31 – 40 | 82 | 27.3 |
| ≥ 41 | 13 | 4.3 |
| Education |  |  |
| Higher education | 82 | 27.3 |
| Lower education | 218 | 72.7 |
| Relationship |  |  |
| Single | 50 | 16.7 |
| Married | 250 | 83.3 |
| Monthly income |  |  |
| Low income | 103 | 34.3 |
| Middle income | 166 | 55.3 |
| High income | 31 | 10.3 |
| Subscription to health insurance |  |  |
| Public | 274 | 91.3 |
| Private | 22 | 7.3 |
| None | 4 | 1.3 |
| Categories of participants |  |  |
| Lactating | 172 | 57.3 |
| Pregnant | 128 | 42.7 |

Some total did not equal 100% due to rounding.

As shown in Figure 1, most participants (∼92%) indicated their willingness to take a brain health test even if it provides information about a disease that cannot be prevented or treated. When asked if they would want to take brain imaging tests, ∼82% and ∼88% indicated they would take EEG and brain MRI, respectively.

**Figure 1.**
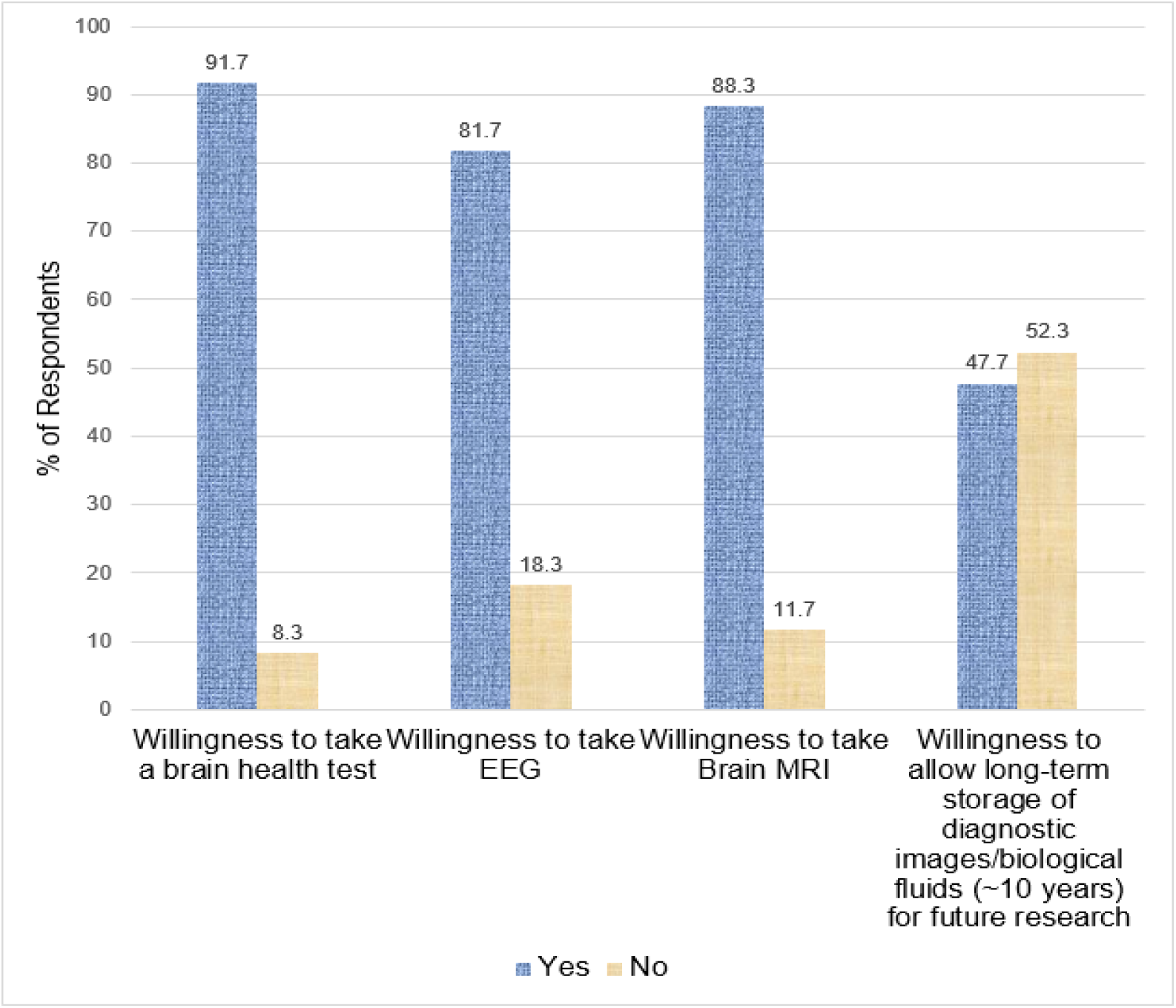
The graph plot shows responses on willingness to take a brain health test, brain imaging tests, and to allow long-term storage of diagnostic images/biological samples.

### Maternal Willingness to take brain imaging and EEG tests

In response to whether participants would take a brain health test even if it provides information about a disease that cannot be prevented or treated, none of the factors predicted willingness.

Being in a partnered relationship is significantly associated with a higher likelihood of wanting to undergo a brain MRI (β = 1.21, *p* = 0.006; OR = 3.35, 95% CI: 1.42–7.92) while being in a low-income bracket is significantly associated with a lower likelihood of wanting to undergo a brain MRI (β = -1.22, p = 0.013; OR = 0.29, 95% CI: 0.11–0.77) (Table 2).

**Table 2.** Sociodemographic Predictors of Maternal Willingness to take brain imaging and EEG tests.

| Predictor Variable | $\beta$ | SE | Wald | $p$-value | OR (95% CI) |
| --- | --- | --- | --- | --- | --- |
| Willingness to take Brain MRI |  |  |  |  |  |
| Relationship status | 1.21 | 0.44 | 7.61 | .006 | 3.35 (1.42, 7.92) |
| Low income (vs Middle income) | -1.22 | 0.48 | 6.21 | 0.013 | 0.29 (0.11, 0.77) |
| Willingness to take EEG |  |  |  |  |  |
| Age: $\geq 41$ years | -1.50 | 0.70 | 4.57 | 0.033 | 0.22 (0.06, 0.88) |
| Relationship status | 0.71 | 0.39 | 3.32 | 0.068 | 2.03 (0.95, 4.33) |
| Low income (vs Middle income) | -1.29 | 0.39 | 11.0 | < 0.001 | 0.27 (0.13, 0.59) |

In examining the willingness to take a brain health test (specifically EEG), women aged ≥41 years (β = -1.50, *p* = 0.033; OR = 0.22, 95% CI: 0.06–0.89) and those with low-income (β = -1.29, *p* < 0.001; OR = 0.27, 95% CI: 0.13–0.59) were less likely to take the test (Table 2). In contrast, those in a relationship ((β = 0.71, *p* = 0.068; OR = 2.03, 95% CI: 0.95–4.33) were more likely to want to take the test, albeit marginally.

### Association between having had previous Foetal/Birth Defect History and Willingness to take Brain Test

Figure 2 shows that only 5.3% indicated they had a history of foetal defect while 7.3% of the participants indicated they had history of birth defect.

**Figure 2.**
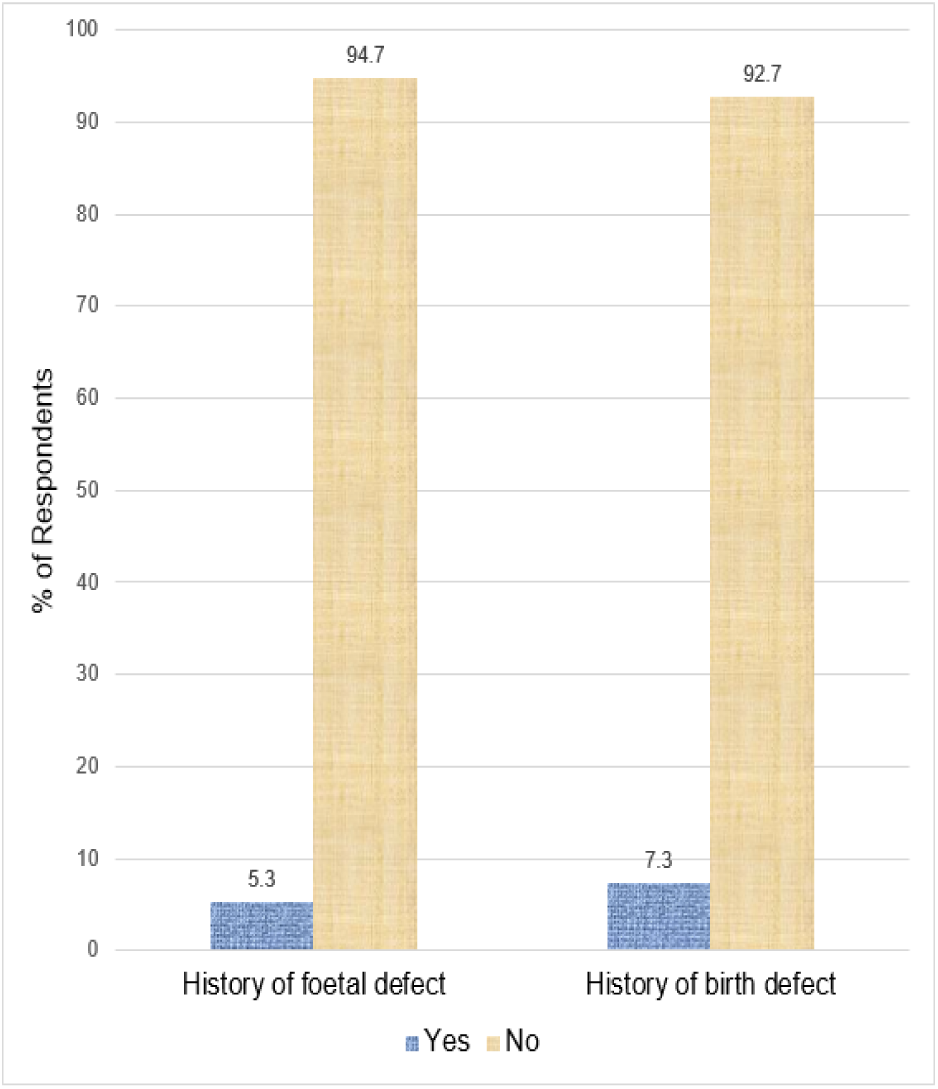
Plot shows maternal history of foetal/birth defect.

We found no statistically significant association between having had a history of foetal defects and willingness to undergo brain health testing even if it provides information about a disease that cannot be prevented or treated (χ^2^ = 0.10, *p* = 0.757), brain MRI (χ^2^ = 0.48, *p* = 0.488) and EEG (χ^2^ = 1.65, *p* = 0.200). In addition, we found no statistically significant association between having had a history of birth defects and willingness to undergo brain health testing even if it provides information about a disease that cannot be prevented or treated (χ^2^ = 0.45, *p* = 0.504) including brain MRI (χ^2^ = 1.17, *p* = 0.280) and EEG ( χ^2^ = 1.35, *p* = 0.244).

### Willingness to provide biological samples

Participants demonstrated high willingness to provide commonly collected biological samples, with 99.7% consenting to blood, 98.3% to stool, and 97.7% to urine. Moderate willingness was observed for breast milk (55.3%), placenta (52%), and amniotic fluid (51%) while lower willingness was reported for less commonly collected samples, such as vaginal fluid (47%), nasal fluid (22.7%), saliva (19.3%), and tears (16.3%).

**Figure 3.**
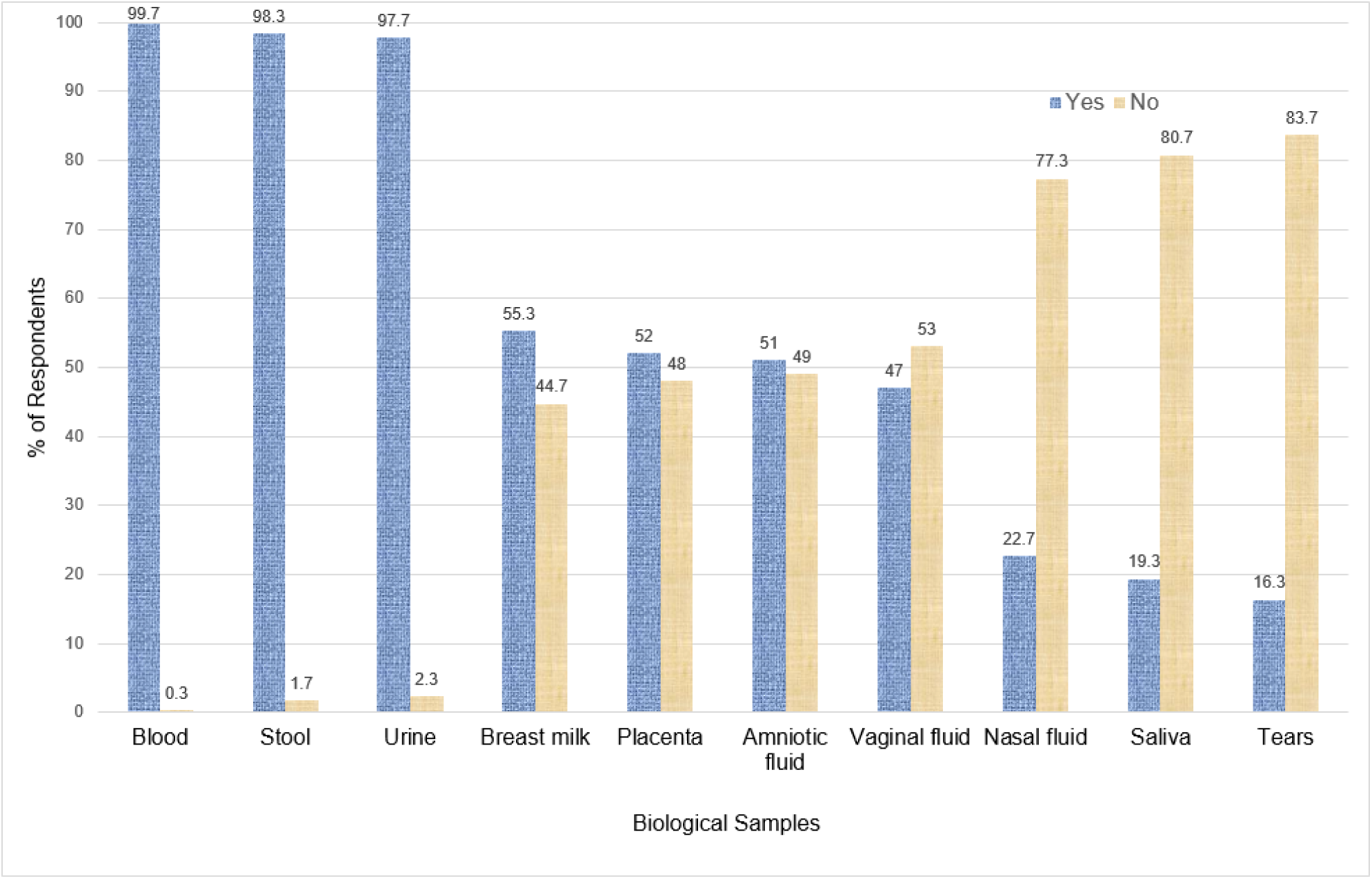
Plot shows willingness to provide biological samples.

Maternal willingness to provide biospecimens varied substantially across specimen types (Table 3). No demographic or socioeconomic predictors were associated with willingness to provide blood. While relationship status and low-income showed marginal positive associations for stool, both were significant predictors of increased willingness to provide urine.

**Table 3.** Sociodemographic Predictors of Maternal Willingness to provide biological samples.

| Predictor Variable | $\beta$ | SE | Wald | p-value | OR (95% CI) |
| --- | --- | --- | --- | --- | --- |
| Stool |  |  |  |  |  |
| Relationship status | 1.88 | 1.08 | 3.03 | 0.082 | 6.56 (0.79, 54.53) |
| Low income (vs Middle income) | 2.42 | 1.32 | 3.37 | 0.067 | 11.19 (0.85, 147.74) |
| Urine |  |  |  |  |  |
| Relationship status | 2.18 | 0.91 | 5.81 | 0.016 | 8.88 (1.50, 52.42) |
| Low income (vs Middle income) | 2.71 | 1.25 | 4.71 | 0.030 | 15.10 (1.30, 175.21) |
| Breast Milk |  |  |  |  |  |
| Higher education | 1.78 | 0.44 | 16.26 | < 0.0001 | 5.94 (2.50, 14.11) |
| Low income (vs Middle income) | -0.80 | 0.31 | 6.45 | 0.011 | 0.45 (0.24, 0.83) |
| Placenta |  |  |  |  |  |
| Relationship status | 0.91 | 0.46 | 3.97 | 0.046 | 2.48 (1.02, 6.07) |
| Higher education | 2.0 | 0.47 | 18.06 | < 0.0001 | 7.35 (2.93, 18.45) |
| Low income (vs Middle income) | -1.27 | 0.33 | 14.53 | < 0.001 | 0.28 (0.15, 0.54) |
| Amniotic fluid |  |  |  |  |  |
| Relationship status | 1.06 | 0.48 | 4.93 | 0.026 | 2.89 (1.13, 7.39) |
| Higher education | 1.95 | 0.46 | 18.27 | < 0.0001 | 7.05 (2.88, 17.28) |
| Low income (vs Middle income) | -1.23 | 0.34 | 13.38 | < 0.001 | 0.29 (0.15, 0.57) |
| Vaginal fluid |  |  |  |  |  |
| Higher education | 1.67 | 0.41 | 16.99 | <0.0001 | 5.32 (2.40, 11.79) |
| Low income (vs Middle income) | -1.52 | 0.36 | 17.86 | < 0.0001 | 0.22 (0.11, 0.44) |
| Nasal fluid |  |  |  |  |  |
| Higher education | 2.15 | 0.40 | 29.02 | < 0.0001 | 8.62 (3.94, 18.87) |
| Lack of health insurance | 2.28 | 1.27 | 3.22 | 0.073 | 9.82 (0.81, 110.27) |
| Saliva |  |  |  |  |  |
| Higher education | 2.13 | 0.42 | 25.73 | < 0.0001 | 8.41 (3.69, 19.16) |
| Lack of health insurance | 2.69 | 1.29 | 4.39 | 0.036 | 14.77 (1.19, 183.52) |
| Tears |  |  |  |  |  |
| Higher education | 2.67 | 0.50 | 29.0 | < 0.0001 | 14.42 (5.46, 38.10) |
| Lack of health insurance | 3.14 | 1.33 | 5.58 | 0.018 | 23.06 (1.71, 311.38) |

Across breastmilk, placenta, amniotic fluid, vaginal fluid, nasal fluid, saliva, and tears, higher maternal education consistently emerged as the strongest positive predictor, but being in a relationship specifically predicted willingness to provide only placenta and amniotic fluid. However, low-income status was consistently associated with reduced willingness to provide these biological samples. Notably, lack of health insurance showed sporadic positive associations for willingness to provide nasal fluid, saliva, and tears.

### Willingness to allow long-term storage of diagnostic images and biological samples

Slightly less than half of the participants (∼48%) would allow long-term storage of their diagnostic images/biological samples for future research (see Figure 1).

In assessing response to the long-term storage of images or biological samples for future research (∼10 years), higher educational attainment was a significant positive predictor (β = 1.47, *p* <0.001, OR = 4.35, 95% CI: 2.04 – 9.24), with mothers possessing higher education levels being over four times more likely to consent (Table 4). Conversely, low-income status was a significant negative predictor (β = - 1.19, *p* <0.001, OR = 0.30, 95% CI: 0.16 – 0.59), with mothers in the low-income group being substantially less likely to consent compared to those with middle income.

**Table 4.** Sociodemographic Predictors of Maternal Willingness to allow long-term storage of diagnostic images and biological samples.

| Predictor Variable | $\beta$ | SE | Wald | $p$-value | OR (95% CI) |
| --- | --- | --- | --- | --- | --- |
| Willingness to allow long-term storage of diagnostic images and biological samples |  |  |  |  |  |
| Higher education | 1.47 | 0.38 | 14.58 | < 0.001 | 4.35 (2.04, 9.24) |
| Low income (vs Middle income) | -1.19 | 0.34 | 12.51 | < 0.001 | 0.30 (0.16, 0.59) |

## Discussion

This cross-sectional study explored maternal interest and consent preferences regarding participation in neurodevelopmental research involving neuroimaging and biological sample collection, critical components for advancing multicentre investigations into *in utero* to early brain development. Our key finding revealed a strong public interest in brain health testing, even when the conditions being assessed are untreatable or cannot be prevented. These findings are discussed below.

### Maternal Interest and Sociodemographic Predictors of Willingness to take brain health test and participate in brain imaging and EEG tests

About 92% of participants expressed willingness to take a brain health test even for untreatable conditions, and over 80% were interested in MRI or EEG. Such openness suggests growing engagement with proactive brain health monitoring, consistent with earlier findings of strong public interest in neurodiagnostic testing [38–43]. Unlike studies linking willingness to demographic or socioeconomic factors [44], our data showed no such associations, pointing to broad, cross-group receptivity. Yet this apparent consensus may conceal anxiety about learning untreatable results, underscoring the need for clearer communication on the purpose and implications of neurodiagnostic testing [42, 43].

When we focused on two specific neuroimaging tests (i.e., brain MRI and EEG), participants in marital relationships were more willing to undergo brain MRI, suggesting that social support can aid decision-making, while lower-income participants were less willing, most likely due to accessibility and awareness issues [45–48]. Interestingly, older age (≥41 years) and low income were associated with reduced EEG willingness; possibly due to limited awareness or procedural apprehension, compounded by the scarcity of EEG and other neurodiagnostic resources in low and middle-income countries (LMICs) [47, 48]. Women in marital relationships showed slightly higher willingness, emphasizing the influence of relational and informational factors.

### Association between Maternal Foetal/Birth Defect History and Willingness to take Brain Test

Only 5.3% and 7.3% reported histories of foetal and birth defects, respectively. These compare with a previous study that reported 7% ultrasound-detected prevalence of foetal defects in Ghana [49] and previous studies documenting a 7.3% self-reported history of birth defects in similar settings [38–41]. These lower self-reported rates likely reflect underdiagnosis, underreporting, and stigma surrounding congenital and neurodevelopmental conditions. Public awareness of neurodevelopmental disorders is necessary for early recognition of these related symptoms and behaviours for early intervention [50]. Importantly, self-reported history of foetal or birth defects was not associated with willingness to undergo brain health testing, including for untreatable conditions. This finding challenges the assumption that prior adverse outcomes necessarily increase receptivity to advanced testing [44]. It instead suggests that emotional burden, perceived futility, and informational barriers may be more influential determinants of testing attitudes [50].

### Maternal Interest and Sociodemographic Predictors of Willingness to provide biological samples

Over 95% of participants were willing to provide common biological samples (blood, stool, urine), reflecting their routine clinical use, familiarity, and perceived low risk. This corroborates our earlier findings [38–41] and that of others [51, 52]. Importantly, maternal willingness to provide blood showed no demographic or socioeconomic associations. This is consistent with its status as a widely accepted, low-risk procedure, with needle phobia and anxiety likely exerting limited influence in this context [35, 53, 54]. Stool and urine provision were also higher than in many previous studies [55, 56]. Though willingness for stool provision showed modest associations with being in a relationship and lower income, urine provision willingness was higher among partnered and lower-income mothers.

By contrast, willingness to provide breast milk, placenta, and amniotic fluid was moderate (about 51–55%), in line with our earlier work [38–41] but lower than some African studies on breast milk donation [57]. Despite the recognised neurodevelopmental value of breast milk [58, 59], reported barriers, such as time constraints, dislike, limited knowledge or supply, infection fears, and cultural or religious beliefs together with concerns about commercialisation and exploitation of low-income women may have contributed to this lower uptake [60, 61].

Higher maternal education and being in a relationship consistently predicted willingness to donate breast milk and placenta, whereas low-income status reduced placenta donation. This raises concerns that specific biospecimen research in LMICs may systematically underrepresent socioeconomically disadvantaged mothers and perpetuate inequities in maternal–child health evidence. Although prior studies have demonstrated the feasibility of obtaining amniotic fluid for prenatal screening of adverse perinatal outcomes [62, 63], our findings indicate moderate willingness to provide this sample, likely reflecting apprehension about its invasive nature. Socioeconomic disparities again influenced participation, with lower-income status associated with reduced willingness.

Willingness to provide less commonly collected samples—vaginal and nasal fluid, saliva, and tears— was lower (approximately 16–47%), echoing our previous findings [38–41]. However, it contrasts with reports of high consent rates for vaginal samples in other African cohorts [36]. Reluctance toward vaginal fluid sampling has been linked to embarrassment, discomfort, and religious or cultural norms [36, 64]. In our study, higher education mitigated this reluctance, whereas financial disadvantage exacerbated it. Further, we found that higher education and lack of health insurance (the later albeit marginally) were associated with increased willingness to provide nasal fluid. This highlights complex motivations in the context of emerging nose-to-brain therapeutics. Although both saliva and tear fluid are promising non-invasive biospecimens with growing roles in neurodevelopmental and proteomic research [65, 66], we found that willingness to provide either of them was driven by higher education and lack of health insurance.

### Sociodemographic Predictors of Maternal Willingness to allow long-term storage of diagnostic images and biological samples

We proposed a 10-year retention period for diagnostic images and biological samples, aligning with Picture Archiving and Communication System retention practices ranging 6 to 240 months [67, 68]. Only 48% of participants consented to long-term storage, consistent with prior work [38–41], reflecting tensions between scientific value and privacy concerns. This may be further compounded by biomarker degradation risks [9, 69] and infrastructural barriers in resource-limited settings. We found that higher maternal education predicted consent to long-term storage of both, consistent with other studies [70, 71] and mirroring broad data-sharing willingness despite reduced control [72, 73]. In contrast, low-income status reduced it, suggesting mistrust, cultural beliefs, or concerns about future data misuse and loss of control, and ethical inequities [74].

It is important to highlight that our study specifically examined long-term storage consent, which differs conceptually from data-sharing preferences and warrants separate exploration, particularly in light of emerging artificial intelligence (AI)-driven research requiring secondary data reuse.

## Strengths and Limitations

To the best of our knowledge, this study is among the first to assess maternal willingness for neurodevelopmental research involving diverse biological samples, imaging modalities, and long-term storage in a low-resource setting, offering insights for ethical recruitment strategies in our setting.

Our study is also not without some limitations. This is a cross-sectional study which focused on a single region and involved only maternal (not paternal) samples, with a modest sample size limiting generalisability. Potential social desirability bias cannot be completely excluded. Finally, participant responses were based on hypothetical willingness, which may differ from decisions made when faced with actual research procedures, especially for more invasive sampling.

## Conclusion and Future Directions

This study reveals high maternal willingness for routine biospecimens (blood, urine, stool) and brain health testing (MRI, EEG), but lower acceptance for sensitive samples (breast milk, placenta, vaginal fluid, tears), with educational level consistently predicting participation and socioeconomic/relational factors shaping uptake. These patterns highlight the need for ethically sensitive, culturally appropriate protocols in low-resource neurodevelopmental research setting. Future multicentre longitudinal studies may consider the integration of parental sociodemographic, biological, and neuroimaging data to elucidate mechanisms linking maternal health and neurodevelopmental outcomes. Ultimately, establishing **c**ontextually grounded ethical frameworks and community engagement strategie**s** will be pivotal in supporting large-scale, sustainable maternal–child brain health research across diverse populations in LMICs.

## Data Availability

The datasets generated and/or analysed during the current study are not publicly available because participants did not provide consent for public data sharing at the time of enrolment. However, data are available from Centre for Education, Population Health Research & Innovation for researchers who meet the criteria for access to confidential data.

## Open Access Statement

For the purpose of open access, the author(s) has applied a Creative Commons Attribution (CC BY) licence to any Author Accepted Manuscript version arising from this submission.

## Declaration

Parts of the findings reported in this manuscript were previously presented at the following scientific meetings: the *2024 European Society of Radiology (ESR) Congress*, Austria; the *2024 York St. John University Conference—“The Only Way Is Ethics! Reflecting on and Learning from Research After Ethical Approval”* (published in the conference proceedings); the *2024 European Society for Magnetic Resonance in Medicine and Biology (ESMRMB) Congress*, Spain; and the *2024 Neonatal Society Autumn Meeting*, United Kingdom.

## References

1. Fitzgerald E, Hor K, Drake AJ. Maternal influences on fetal brain development: The role of nutrition, infection and stress, and the potential for intergenerational consequences. Early Human Development. 2020;150:105190.

2. Wu Y, De Asis-Cruz J, Limperopoulos C. Brain structural and functional outcomes in the offspring of women experiencing psychological distress during pregnancy. Molecular Psychiatry. 2024;29(7):2223–40.

3. Cainelli E, Vedovelli L, Bisiacchi P. The mother–child interface: A neurobiological metamorphosis. Neuroscience 2024; 561:92–106.

4. Nolvi S, Merz EC, Kataja EL, Parsons CE. Prenatal stress and the developing Brain: Postnatal environments Promoting Resilience. Biological Psychiatry 2022;93(10):942–52.

5. Lubrano C, Parisi F, Cetin I. Impact of maternal environment and inflammation on fetal neurodevelopment. Antioxidants 2024;13(4):453.

6. Martini S, Lenzi J, Paoletti V, Maffei M, Toni F, Fetta A, Aceti A, Cordelli DM, Zuccarini M, Guarini A, Sansavini A, Corvaglia L. Neurodevelopmental Correlates of Brain Magnetic Resonance Imaging Abnormalities in Extremely Low-birth-weight Infants. J Pediatr. 2023;262:113646.

7. Wilkinson CL, Yankowitz LD, Chao JY, Gutiérrez R, Rhoades JL, Shinnar S, Purdon PL, Nelson CA. Developmental trajectories of EEG aperiodic and periodic components in children 2-44 months of age. Nat Commun. 2024;15(1):5788.

8. Goodspeed K, Armstrong D, Dolce A, Evans P, Said R, Tsai P, Sirsi D. Electroencephalographic (EEG) Biomarkers in Genetic Neurodevelopmental Disorders. J Child Neurol. 2023;38(6-7):466–477.

9. Wang Y, Li X. Editorial: Eye movement tracking in ocular, neurological, and mental diseases. Front Neurosci. 2024;18:1364078. doi: 10.3389/fnins.2024.1364078. PMID: 38312928; PMCID: PMC10834791.

10. Yamamoto M, Eguchi A, Sakurai K, Nakayama SF, Sekiyama M, Mori C, et al. Longitudinal analyses of maternal and cord blood manganese levels and neurodevelopment in children up to 3 years of age: The Japan Environment and Children’s Study (JECS). Environment International. 2022;161:107126.

11. Lee ASE, Ji Y, Raghavan R, Wang G, Hong X, Pearson C, Mirolli G, Bind E, Steffens A, Mukherjee J, Haltmeier D, Fan ZT, Wang X. Maternal prenatal selenium levels and child risk of neurodevelopmental disorders: A prospective birth cohort study. Autism Res. 2021;14(12):2533–2543.

12. Sun Z, Lee-Sarwar K, Kelly RS, Lasky-Su JA, Litonjua AA, Weiss ST, Liu YY. Revealing the importance of prenatal gut microbiome in offspring neurodevelopment in humans. EBioMedicine. 2023;90:104491. doi: 10.1016/j.ebiom.2023.104491.

13. Di Gesù CM, Matz LM, Buffington SA. Diet-induced dysbiosis of the maternal gut microbiome in early life programming of neurodevelopmental disorders. Neurosci Res. 2021;168:3–19. doi: 10.1016/j.neures.2021.05.003.

14. Wang X, Luo ZC, Du O, Zhang HJ, Fan P, Ma R, et al. The association between maternal urinary Bisphenol A levels and neurodevelopment at age 2 years in Chinese boys and girls: A prospective cohort study. Ecotoxicology and Environmental Safety. 2023;264:115413. 10.1016/j.ecoenv.2023.115413

15. Xie Y, Xiao H, Zheng D, Mahai G, Li Y, Xia W, et al. Associations of prenatal metal exposure with child neurodevelopment and mediation by perturbation of metabolic pathways. Nature Communications. 2025;16(1). 10.1038/s41467-025-57253-3

16. Brown Belfort M. The Science of Breastfeeding and Brain Development. Breastfeed Med. 2017;12(8):459–461. doi: 10.1089/bfm.2017.0122.

17. Heng YY, Asad I, Coleman B, Menard L, Benki-Nugent S, Hussein Were F, Karr CJ, McHenry MS. Heavy metals and neurodevelopment of children in low and middle-income countries: A systematic review. PLoS One. 2022; 31;17(3):e0265536. doi: 10.1371/journal.pone.0265536.

18. Baron-Cohen S, Tsompanidis A, Auyeung B, Nørgaard-Pedersen B, Hougaard DM, Abdallah M, et al. Foetal oestrogens and autism. Molecular Psychiatry. 2019;25(11):2970–8.

19. Thornburg KL, O’Tierney PF, Louey S. Review: The Placenta is a Programming Agent for Cardiovascular Disease. Placenta. 2010;31:S54–9.

20. Shibata T, Takata E, Takakura M, Han J, Yamada S, Satoh T. Amniotic fluid as a novel delivery route of 3-hydroxybutyrate during fetal brain development. Eur J Obstet Gynecol Reprod Biol. 2025;311:114001. doi: 10.1016/j.ejogrb.2025.114001.

21. Buono R, Longo VD. Starvation, Stress Resistance, and Cancer. Trends Endocrinol Metab. 2018;29(4):271-280. doi: 10.1016/j.tem.2018.01.008.

22. Dubé-Zinatelli E, Cappelletti L, Ismail N. Vaginal Microbiome: Environmental, Biological, and Racial Influences on Gynecological Health Across the Lifespan. Am J Reprod Immunol. 2024;92(6):e70026. doi: 10.1111/aji.70026.

23. Zhou L, Qiu W, Wang J, Zhao A, Zhou C, Sun T, et al. Effects of vaginal microbiota transfer on the neurodevelopment and microbiome of cesarean-born infants: A blinded randomized controlled trial. Cell Host & Microbe 2023;31(7):1232–1247.e5.

24. Mehta NH, Sherbansky J, Kamer AR, Carare RO, Butler T, Rusinek H, Chiang GC, Li Y, Strauss S, Saint-Louis LA, Theise ND, Suss RA, Blennow K, Kaplitt M, de Leon MJ. The Brain-Nose Interface: A Potential Cerebrospinal Fluid Clearance Site in Humans. Front Physiol. 2022;12:769948. doi: 10.3389/fphys.2021.769948.

25. Parker KJ, Oztan O, Libove RA, Mohsin N, Karhson DS, Sumiyoshi RD, Summers JE, Hinman KE, Motonaga KS, Phillips JM, Carson DS, Fung LK, Garner JP, Hardan AY. A randomized placebo-controlled pilot trial shows that intranasal vasopressin improves social deficits in children with autism. Sci Transl Med. 2019;11(491):eaau7356. doi: 10.1126/scitranslmed.aau7356.

26. Tartaglia G, Connelly S. Editorial: Saliva used as biological fluid to detect neurodegenerative and neurodevelopmental diseases. Front Neurosci. 2023;17:1141376. doi: 10.3389/fnins.2023.1141376.

27. Janšáková K, Kyselicová K, Ostatníková D, Repiská G. Potential of salivary biomarkers in autism Research: a Systematic review. International Journal of Molecular Sciences 2021;22(19):10873.

28. Börger M, Funke S, Bähr M, Grus F, Lingor P. Biomarker sources for Parkinson’s disease: Time to shed tears? Basal Ganglia 2015;5(2–3):63–9.

29. Luib E, Demleitner AF, Cordts I, Westenberg E, Rau P, Pürner D, Haller B, Lingor P. Reduced tear fluid production in neurological diseases: a cohort study in 708 patients. J Neurol. 2024; 271(4):1824–1836. doi: 10.1007/s00415-023-12104-3.

30. Tursic A, Vaessen M, Zhan M, Vingerhoets AJJM, De Gelder B. The power of tears: Observers’ brain responses show that tears provide unambiguous signals independent of scene context. Neuroimage Reports. 2022;2(3):100105.

31. Riem MME, Van IJzendoorn MH, De Carli P, Vingerhoets AJJM, Bakermans-Kranenburg MJ. As tears go by: Baby tears trigger more brain activity than adult tears in nulliparous women. Social Neuroscience. 2016;1–4.

32. Bebell LM, Ngonzi J, Meier FA, Carreon CK, Birungi A, Kerry VB, Atwine R, Roberts DJ. Building Perinatal Pathology Research Capacity in Sub-Saharan Africa. Front Med (Lausanne). 2022; 9:958840.

33. Mahdikhani Z, Jahanian Sadatmahalleh S, Alimoradi Z, Habibelahi A. Explaining the experiences of donors, recipients, and healthcare providers regarding milk donation: a systematic review and meta-synthesis. Int Breastfeed J. 2025;20(1):50. doi: 10.1186/s13006-025-00740-6.

34. Awad R, Avital A, Sosnik A. Polymeric nanocarriers for nose-to-brain drug delivery in neurodegenerative diseases and neurodevelopmental disorders. Acta Pharmaceutica Sinica B 2022;13(5):1866–86.

35. Jaswal H, Ialomiteanu A, Hamilton H, Rehm J, Wells S, Shield KD. Willingness of population health survey participants to provide personal health information and biological samples. BMC Public Health. 2024;24(1):3279. doi: 10.1186/s12889-024-20769-2. PMID: 39593030; PMCID: PMC1159

36. Claassen-Weitz S, Kullin B, du Toit E, Gardner-Lubbe S, Passmore JS, Jaspan H, Happel AU, Bellairs G, Hilton C, Chicken A, Welp K, Livingstone H, Brink A. Knowledge and perceptions of blood donors of the Western Cape Blood Services, South Africa, toward vaginal sample donation for biobanking. Front Reprod Health. 2024a; 6:1446809. doi: 10.3389/frph.2024.1446809. PMID: 39665033; PMCID: PMC11631888.

37. Carver RB, Fredheim NAG, Mowinckel AM, Ebmeier KP, Friedman BB, Rosness TA, Drevon CA, Suri S, Baaré WFC, Zsoldos E, Solé-Padullés C, Bartrés-Faz D, Ghisletta P, Nawijn L, Düzel S, Madsen KS, Fjell AM, Lindenberger U, Walhovd KB, Budin-Ljøsne I. People’s interest in brain health testing: Findings from an international, online cross-sectional survey. Front Public Health. 2022;10:998302. doi: 10.3389/fpubh.2022.998302.

38. Piersson AD, Amartey C, Quartei ST, Mumuni HN, Dzefi-Tettey K, Jones L. Maternal willingness and ethical considerations for child participation in neurodevelopmental research in a low-resource setting. Oral Presentation. Neonatal Society Autumn Meeting; 2024 Nov 14; Royal Society of Medicine, London, UK.

39. Piersson AD, Amartey C, Arkorful J, Dzefi-Tettey K. Maternal-child interest in neuroimaging research and collection of human biological materials in an urban low-resource setting. Magn Reson Mater Phy. 2024;37(Suppl 1):S444–S445. doi:10.1007/s10334-024-01191-6. E-Poster.

40. Piersson AD. Maternal-child ethical concerns in neurodevelopmental research involving neuroimaging and collection of biological samples: a tale of two urban cities in a low-resource setting. Oral Presentation. Proceedings of York St John University Conference “The Only Way Is Ethics! Reflecting on and learning from research after ethical approval”; 2024 Mar 22; York St John University, York, UK.

41. Piersson AD, Buabeng E. Maternal-child interest in neurodevelopmental research involving neuroimaging and collection of biological samples in a low-resource setting. European Congress of Radiology. 2024 Feb 28-Mar 3; Vienna, Austria. doi:10.26044/ecr2024/C-14062. E-Poster.

42. Milne R, Diaz A, Badger S, Bunnik E, Fauria K, Wells K. At, with and beyond risk: expectations of living with the possibility of future dementia. Sociol Health Illn. (2018) 40:969–87.

43. Friedman BB, Suri S, Solé-Padullés C, Düzel S, Drevon CA, Baaré WFC, et al. Are people ready for personalized brain health? Perspectives of research participants in the LifeBrain Consortium. The Gerontologist. 2019;60(6):1050–9. 10.1093/geront/gnz155

44. Carver RB, Fredheim NAG, Budin-Ljosne I, Friedman BB. What Motivates People to Look After Their Brain Health? Insights From the Global Brain Health Survey. University of Oslo (2022).

45. Bunnik EM, Vernooij MW. Incidental findings in population imaging revisited. European Journal of Epidemiology. 2016;31(1):1–4.

46. Bomhof CHC, VAN Bodegom L, Vernooij MW, Pinxten W, DE Beaufort ID, Bunnik EM. The Impact of Incidental Findings Detected During Brain Imaging on Research Participants of the Rotterdam Study: An Interview Study. Camb Q Healthc Ethics. 2020; 29(4):542–556. doi: 10.1017/S0963180120000304.

47. McLane HC, Berkowitz AL, Patenaude BN, McKenzie ED, Wolper E, Wahlster S, Fink G, Mateen FJ. Availability, accessibility, and affordability of neurodiagnostic tests in 37 countries. Neurology. 2015; 85(18):1614–22. doi: 10.1212/WNL.0000000000002090.

48. Piersson AD, Gorleku PN. Assessment of availability, accessibility, and affordability of magnetic resonance imaging services in Ghana. Radiography (Lond). 2017; 23(4):e75–e79. doi: 10.1016/j.radi.2017.06.002.

49. Swarray-Deen, A., Yapundich, M., Boudova, S. et al. Spectrum of congenital anomalies detected through anatomy ultrasound at a referral hospital in Ghana. BMC Pregnancy Childbirth. 2025; 25(1):500. 10.1186/s12884-025-07640-x

50. Piersson AD, Amlalo JG, Dzefi-Tettey K. Public awareness of neurodevelopmental disorders and affordability of neuroimaging tools for clinical assessment of early childhood brain assessment in a low-resource setting. Magn Reson Mater Phy. 2024;37(Suppl 1):S341–S343. doi:10.1007/s10334-024-01191-6.

51. Bone JN, Pickerill K, Kinshella MLW, Vidler M, Craik R, Poston L, et al. Pregnancy cohorts and biobanking in sub-Saharan Africa: a systematic review. BMJ Global Health. 2020;5(11):e003716

52. Abdul-Rahman OA, Rodriguez B, Wadlinger SR, Slutsman J, Boyle EB, Merrill LS, Botkin J, Moye J Jr. Success rates for consent and collection of prenatal biological specimens in an epidemiologic survey of child health. Birth Defects Res A Clin Mol Teratol. 2016;106(1):47–54. doi: 10.1002/bdra.23455. Epub 2015 Sep 26. PMID: 26407522; PMCID: PMC4718757.

53. Pronicki Ł, Czech M, Gujski M, Boguszewska ND. Awareness, Attitudes and Willingness to Donate Biological Samples to a Biobank: A Survey of a Representative Sample of Polish Citizens. Healthcare (Basel). 2023;11(20):2714. doi: 10.3390/healthcare11202714.

54. Kocic N, Bujandric N, Budakov Obradovic Z, Grujic J, Bezanovic M, Kolarovic J. Factors influencing blood donation among university students in Vojvodina, Serbia: cross-sectional study. BMJ Open. 2024;14(11):e086700. doi: 10.1136/bmjopen-2024-086700.

55. Bolte LA, Klaassen MAY, Collij V, Vich Vila A, Fu J, van der Meulen TA, de Haan JJ, Versteegen GJ, Dotinga A, Zhernakova A, Wijmenga C, Weersma RK, Imhann F. Patient attitudes towards faecal sampling for gut microbiome studies and clinical care reveal positive engagement and room for improvement. PLoS One. 2021;16(4):e0249405. doi: 10.1371/journal.pone.0249405.

56. Davies R, Iturriza-Gómara M, Glennon-Alty R, Elliot AJ, Vivancos R, Alvarez Nishio A, Cunliffe NA, Hungerford D. Public acceptability of a technology-mediated stool sample collection platform to inform community-based surveillance of infectious intestinal disease: a pilot study. BMC Public Health. 2022;22(1):958. doi: 10.1186/s12889-022-13307-5.

57. Ahmed MAM, Namisi CP, Kirabira NV, Lwetabe MW, Rujumba J. Acceptability to donate human milk among postnatal mothers at St. Francis hospital Nsambya, Uganda: a mixed method study. Int Breastfeed J. 2024 Feb 1;19(1):9. doi: 10.1186/s13006-024-00615-2.

58. Belfort MB, Knight E, Chandarana S, Ikem E, Gould JF, Collins CT, et al. Associations of maternal milk feeding with neurodevelopmental outcomes at 7 years of age in former preterm infants. JAMA Network Open 2022;5(7):e2221608.

59. Gao Y, Lu X, Pan M, Liu C, Min Y, Chen X. Effect of breast milk intake volume on early behavioral neurodevelopment of extremely preterm infants. International Breastfeeding Journal. 2024;19(1).

60. Mampane T, Wolvaardt JE. The acceptability of a donor human milk bank and donated human milk among mothers in Limpopo Province, South Africa. Matern Child Nutr. 2024;20(3):e13651. doi: 10.1111/mcn.13651.

61. Smith JP. Markets, breastfeeding and trade in mothers’ milk. International Breastfeeding Journal. 2015;10(1).

62. Geer LA, Pycke BFG, Sherer DM, Abulafia O, Halden RU. Use of amniotic fluid for determining pregnancies at risk of preterm birth and for studying diseases of potential environmental etiology. Environmental Research. 2014;136:470–81. 10.1016/j.envres.2014.09.031

63. Liu X, Quan S, Fu Y, Wang W, Zhang W, Wang X, et al. Study on amniotic fluid metabolism in the second trimester of Trisomy 21. Journal of Clinical Laboratory Analysis. 2019;34(3):e23089. Available from: 10.1002/jcla.23089

64. Kutalek R, Baingana F, Sevalie S, Broutet N, Thorson A. Perceptions on the collection of body fluids for research on persistence of Ebola virus: A qualitative study. PLoS Neglected Tropical Diseases [Internet]. 2020;14(5):e0008327. 10.1371/journal.pntd.0008327

65. Beversdorf DQ, Sohl K, Levitskiy D, Tennant P, Goin-Kochel RP, Shaffer RC, Confair A, Middleton FA, Hicks SD. Saliva RNA Biomarkers of Gastrointestinal Dysfunction in Children With Autism and Neurodevelopmental Disorders: Potential Implications for Precision Medicine. Front Psychiatry. 2022;12:824933.

66. Barmada A, Shippy SA. Tear analysis as the next routine body fluid test. Eye (Lond). 2020;34(10):1731–1733. doi: 10.1038/s41433-020-0930-0.

67. Behnam F, Khajouei R, Ahmadian L. The retention duration of digital images in picture archiving and communication systems. Heliyon. 2024;10(6):e27847.

68. Im K, Gui D, Yong WH. An introduction to hardware, software, and other information technology needs of biomedical biobanks. Biobanking: methods and protocols. 2019;17–29

69. Kleeberger C, Shore D, Gunter E, Sandler DP, Weinberg CR. The Effects of Long-term Storage on Commonly Measured Serum Analyte Levels. Epidemiology. 2018;29(3):448–452. doi: 10.1097/EDE.0000000000000810.

70. Nansumba H, Flaviano M, Patrick S, Isaac S, Wassenaar D. Health care users’ acceptance of broad consent for storage of biological materials and associated data for research purposes in Uganda. Wellcome Open Res. 2022;7:73. doi: 10.12688/wellcomeopenres.17633.2.

71. Ly S, Reyes-Hadsall S, Drake L, Zhou G, Nelson C, Barbieri JS, Mostaghimi A. Public Perceptions, Factors, and Incentives Influencing Patient Willingness to Share Clinical Images for Artificial Intelligence-Based Healthcare Tools. Dermatol Ther (Heidelb). 2023;13(11):2895–2902. doi: 10.1007/s13555-023-01031-w.

72. Köngeter A, Schickhardt C, Jungkunz M, Bergbold S, Mehlis K, Winkler EC. Patients’ Willingness to Provide Their Clinical Data for Research Purposes and Acceptance of Different Consent Models: Findings From a Representative Survey of Patients With Cancer. J Med Internet Res. 2022;24(8):e37665. doi: 10.2196/37665.

73. Loffredo CA, Luta G, Wallington S, Makgoeng SB, Selsky C, Mandelblatt JS, Adams-Campbell LL; Region 1 Bio-specimen Management of Cancer Health Disparities Program. Knowledge and willingness to provide research biospecimens among foreign-born Latinos using safety-net clinics. J Community Health. 2013;38(4):652–9. doi: 10.1007/s10900-013-9660-6.

74. Moodley K, Sibanda N, February K, Rossouw T. “It’s my blood”: ethical complexities in the use, storage and export of biological samples: perspectives from South African research participants. BMC Medical Ethics. 2014;15(1).

